# Exploring the Transmission Dynamics of the COVID-19 Outbreaks after Dec. 2022 in Shaanxi Province, China: Analysis of Baseline Data from A Large Scale Cohort

**DOI:** 10.1101/2023.01.24.23284952

**Authors:** Tianxiao Zhang, Baibing Mi, Mingwang Shen, Yunpeng Nian, Suixia Cao, Jingchun Liu, Hao Huang, Zhongxi Wei, Lixi Liu, Qian Wu, Yi Zhang, Shaobai Zhang

## Abstract

**Backgrounds:** The goal of this study is to explore the transmission dynamics for recent large-scale COVID-19 outbreaks in Shaanxi Province on the Chinese mainland. Furthermore, the potential effects of the Spring Festival travel rush on the ongoing COVID-19 pandemic were depicted.

**Methods:** This study uses baseline data from a large cohort to investigate the characteristics of the recent COVID-19 epidemic in Shaanxi province. A cluster sampling method was used to recruit the study participants during the COVID-19 pandemic in Shaanxi province since Dec. 1^st^, 2022. A total of 44 sampling cluster (11 village in rural areas and 33 residences in urban areas) were chosen for enrollment of study participants. A self-developed questionnaire was applied to data collection of socio-demographic and COVID-19 pandemic related information.

**Results:** A total of 14,744 study participants were enrolled in the baseline survey and 12,111 completed survey data were extracted for analysis. The cumulative infection attack rate (IAR) of COVID-19 among the study participants was 84.7%. The cumulative IAR in urban and rural areas were 85.6% and 83.7%, respectively. A peak of COVID-19 self-reported diagnosis could be observed from Dec. 15^th^, 2022 to Jan. 1^st^, 2023 in the provincial level. Beside this major peak of the recent epidemic (around Dec.20^th^, 2022), a small but steep rise could also be identified between Jan 13^th^ to 14^th^, 2023. Individuals who escaped the first wave of COVID-19 outbreaks may face danger of infection from returnees during the 2023 Spring Festival.

**Conclusion:** According to the COVID-19 cumulative IAR data, the herd community was primarily achieved in Shaanxi province’s urban and rural areas. The epidemic in Shaanxi province has been exacerbated by mass population movement during the Spring Festival travel rush in both urban and rural areas. Further surveillance should be performed to monitor the spread of SARS-CoV-2 infections.

## Introduction

On November 11 and December 7, 2022, China announced several new measures to optimize the implementation of novel corona-virus disease 2019 (COVID-19) epidemic prevention and control [1-2]. Since then, Omicron infections have spread rapidly across mainland China. However, it is difficult to accurately assess the transmission dynamics of the COVID-19 outbreaks because mass testing and intensive contact tracing are rarely performed after new prevention and control strategies are implemented. Therefore, it is urgent to carry out population-level COVID-19 surveillance cohort to depict the transmission dynamics patterns of the epidemics, and examine the differences of transmission patterns in the urban and rural areas, so as to put forward more precise prevention and control measures. In addition, the Spring Festival (Jan. 21^st^, 2023) and its relevant travel rush (Jan. 7^th^ – Feb. 15^th^) are in progress. The potential effects of this large-scale travel rush in China on the on-going epidemic are largely unknown. Up to date, several large-scale population based surveys have been proposed and implemented by different levels of Health Commissions to answer these questions. Leung *et al*. fitted a dynamic model to estimate the transmission dynamics of SARS-CoV-2 Omicron BF.7 in Beijing after the adjustment of the zero-COVID policy [3]. However few studies have focused on other part of China.

Shaanxi province is landlocked and lies in the middle of China, covering an area of 205,600 km2 with a population of 39.2 million, ranking the 16th highest among the provincial-level administrations in China (China Census Data, 2020). Shaanxi province consists of 10 prefecture-level cities. Xi’an is the provincial capital as well as the largest city in Middle of Shaanxi Province. The other prefecture-level cities into which the province is divided are Xianyang, Ankang, Baoji, Hanzhong, Shangluo, Tongchuan, Weinan, Yan’an and Yulin.

In the present study, we retrospectively constructed the transmission dynamics curves of the recent large-scale COVID-19 outbreaks in the Shaanxi province of China mainland. In addition, we aimed to depict the potential effects of the Spring Festival travel rush on this on-going COVID-19 pandemic. The data utilized in this study is based on the baseline survey from a large-scale cohort aiming to investigate the features of this recent COVID-19 epidemic in Shaanxi province. A total of 44 villages/communities from 10 cities in Shaanxi province were selected for the survey, hoping to understand the epidemic trends of COVID-19 among urban and rural populations and provide a basis for the country to formulate future relevant strategies.

## Methods

### Study Design and Data Collection

This study is based on the baseline data from a large-scale cohort aiming to investigate the features of this recent COVID-19 epidemic in Shaanxi province. A cluster sampling method was used to recruit the study participants during the COVID-19 pandemic in Shaanxi province. The baseline survey was conducted between Jan. 12^th^ – 16^th^ of 2023. To accurately estimate the provincial-level cumulative infection attack rate (IAR) based on sample data, we performed power analysis using Sample Size Calculator (https://riskcalc.org/samplesize/) to determine the minimum number of study participants [4]. To achieve the confidence limits of 5% and consider relevant sample loss, a minimum sample of 1,500 participants is required.

A total of 44 sampling clusters (11 villages in rural and 33 residences in urban) were chosen to enroll participants according to population size, stability, representativeness and local commitment and capacity (Supplementary Figure S1). With the assistance of the Shaanxi Provincial Center for Disease Control and Prevention, the researchers contacted and received permission and assistance from local health committees and disease control centers to dispatch trained investigators to conduct household face-to-face surveys in the above clusters. The Research Electronic Data Capture (REDCap) was used to collect data [5-7]. If household visits were inconvenient, a telephone survey was conducted with village doctors and community workers or through an online survey link was sent to the potential respondents by researcher using WeChat/QQ groups dropped. After scanning the QR code on the invitation letter, the invitees were directed to the online questionnaire. After completing the questionnaire, the REDCap automatically collected and coded the responses. Each participant could only fill in the online questionnaire once as controlled by the individual’s IP address. Invalid questionnaires with missing answers, incorrect information, or choosing the exact solution for all items were excluded. Two researchers checked all data independently, and a third researcher was involved in resolving any difference in the interpretation between the two researchers. A self-developed socio-demographic and COVID-19 pandemic related factors questionnaire was utilized to collect information including gender, age, family size, whether participants had been infected during the COVID-19 pandemic, etc. A full copy of the questionnaire can be found in supplementary materials. The study participants are fully informed the relevant information of this study before they start to complete the questionnaire. This data generated in this study are properly anonymized to minimize the risk of leaking personal sensitive information. This study is approved by the institutional review board of Xi’an Jiaotong University Health Science Center.

### Statistical Analysis

The Student’s t-test and χ^2^ test were utilized for significance tests on continuous and categorical data, respectively. The following choices recorded the self-reported date of diagnosis for COVID-19 patients: 1) the date of typical symptom onset (68%), 2) the date of positive for a rapid agent test when information (16%), or 3) the date of positive for a RT-PCR test (16%). Statistical computing software R v4.2.2 and relevant packages were utilized for statistical analysis and visualization of geographical distributions of survey locations [8].

## Results

### Characteristics of the study participants

A total of 14,744 study participants were enrolled in the baseline survey and data of 12,111 study participants who completed survey data were extracted for analysis (Supplementary Figure S2). The average age of the study participants were 41.5 and 44.5 years-old in urban and rural areas, respectively. The pyramid graph depicting age distribution of the study participants were showed in Figure 1. The study participants were comprised of 5,826 males (48%) and 6,285 females (52%). Among these study participants, 5,910 individuals (49%) reside in the urban areas while 6,201 individuals (51%) reside in the rural areas. The geographical distributions of the study participants recruited in Shaanxi province were summarized in Figure 2.

**Figure 1.**
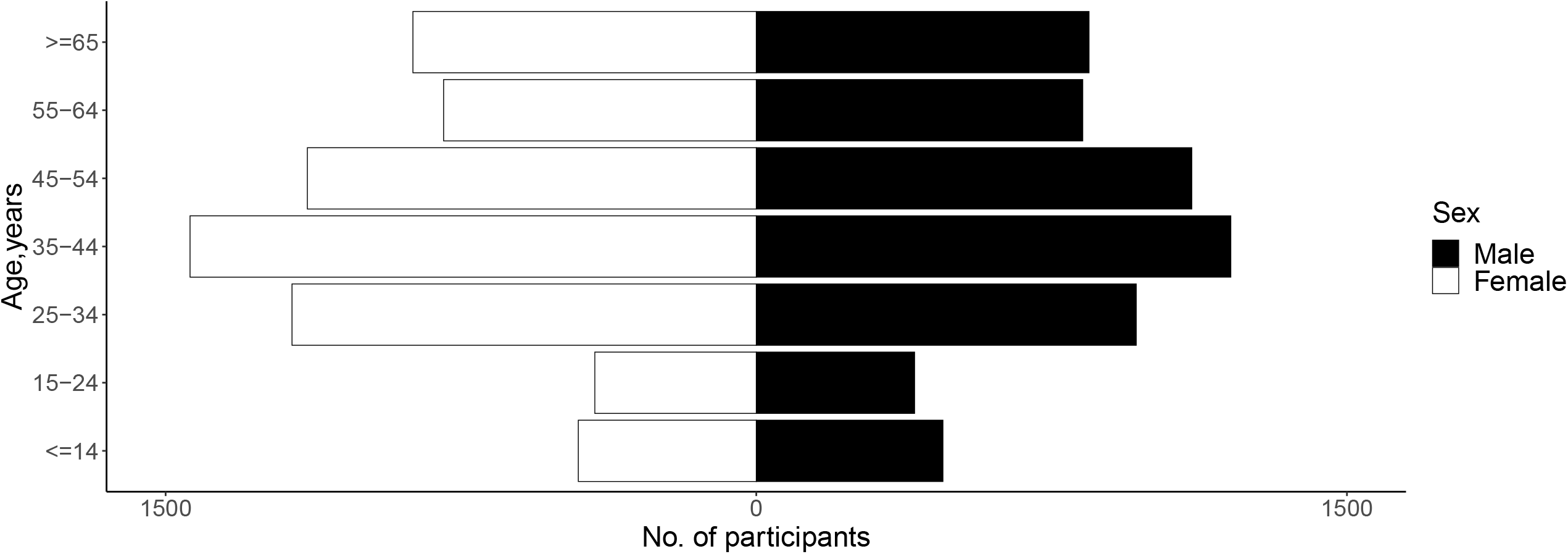
The age-sex pyramid graph of the study participants.

**Figure 2.**
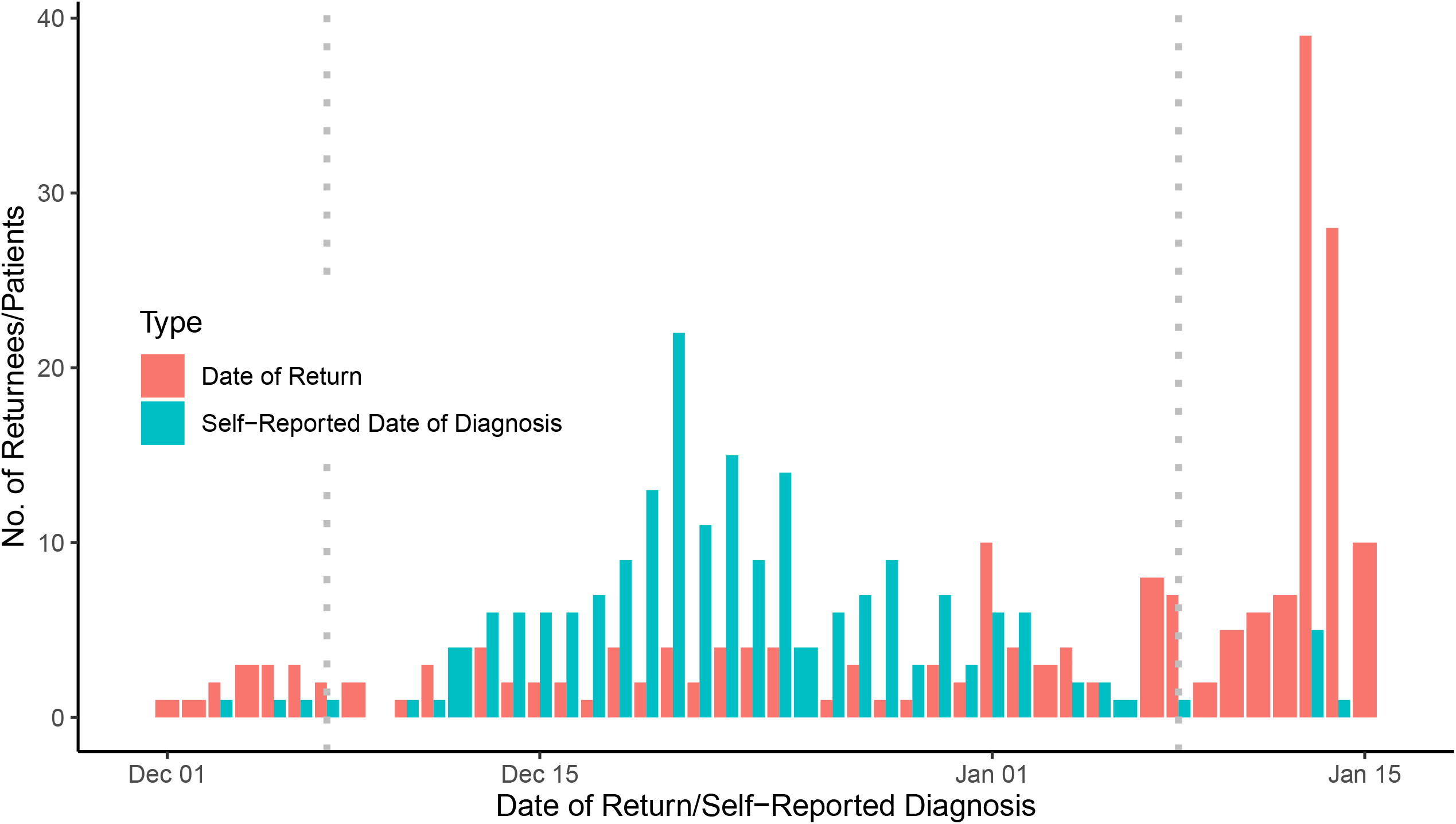
The geographic distributions of the cumulative infection attack rate in Shaanxi province. A. Urban areas. B. Rural areas

### The proportion of COVID-19 infected persons in the study participants

The cumulative infection attack rate (IAR, proportion of the infected persons) of COVID-19 among the study participants was 84.7% based on the present data. Slight but significant difference (χ^2^=8.45, *P*=0.004) was identified between residents of urban (85.6%) and rural areas (83.7%) (Supplementary Table S1). To further investigate the geographical effects on the cumulative infection attack rate, we extracted a subset of residential addresses from the rural areas which were ≥10km (named “remote areas”) from the nearby urban areas. The cumulative IAR in urban, rural, and remote areas were summarized in Table 1. The highest and lowest cumulative IAR were identified in general rural areas (92.9%) and remote areas (76.0%).

**Table 1.** Cumulative infection attach rate in types of residential address in Shaanxi province.

| Areas | Infected<br>(N=10,253) | Study Participants<br>(N=12,111) | Cumulative IAR,% |
| --- | --- | --- | --- |
| Urban Area | 5,060 | 5,910 | 85.6 |
| Rural Area | 2,643 | 2,845 | 92.9 |
| Remote Area | 2,550 | 3,356 | 76.0 |

IAR: infection attack rate.
\*Remote Area is defined by rural areas which are of $\geq 10$ km from nearby urban area.

### Transmission patterns of COVID-19 outbreaks in Shaanxi Province

The transmission curves of COVID-19 epidemic in Shaanxi Province were constructed based on the participants self-reported date of diagnosis (Figure 3). A peak of COVID-19 self-reported diagnosis could be observed from Dec. 15^th^, 2022 to Jan. 1^st^, 2023 at provincial level (Figure 3A). Further analyses were performed to examine the transmission patterns through grouping the patients by their types of residential addresses (Figure 3B). Compared to the rural areas, the transmission of the COVID-19 epidemic in the urban areas has a relatively steep rise and decline. On the other hand, the peak of the outbreaks in rural areas is a plateau which lasts for about 15 days. Interestingly, besides the major peak of the recent epidemic (around Dec.20^th^, 2022), a small but steep rise could also be identified between Jan 13^th^ to 14^th^, 2023. The geographical distributions of the patients comprised of this small rise were showed in Supplementary table S2. These patients were identified in multiple cities from both urban and rural areas.

**Figure 3.**
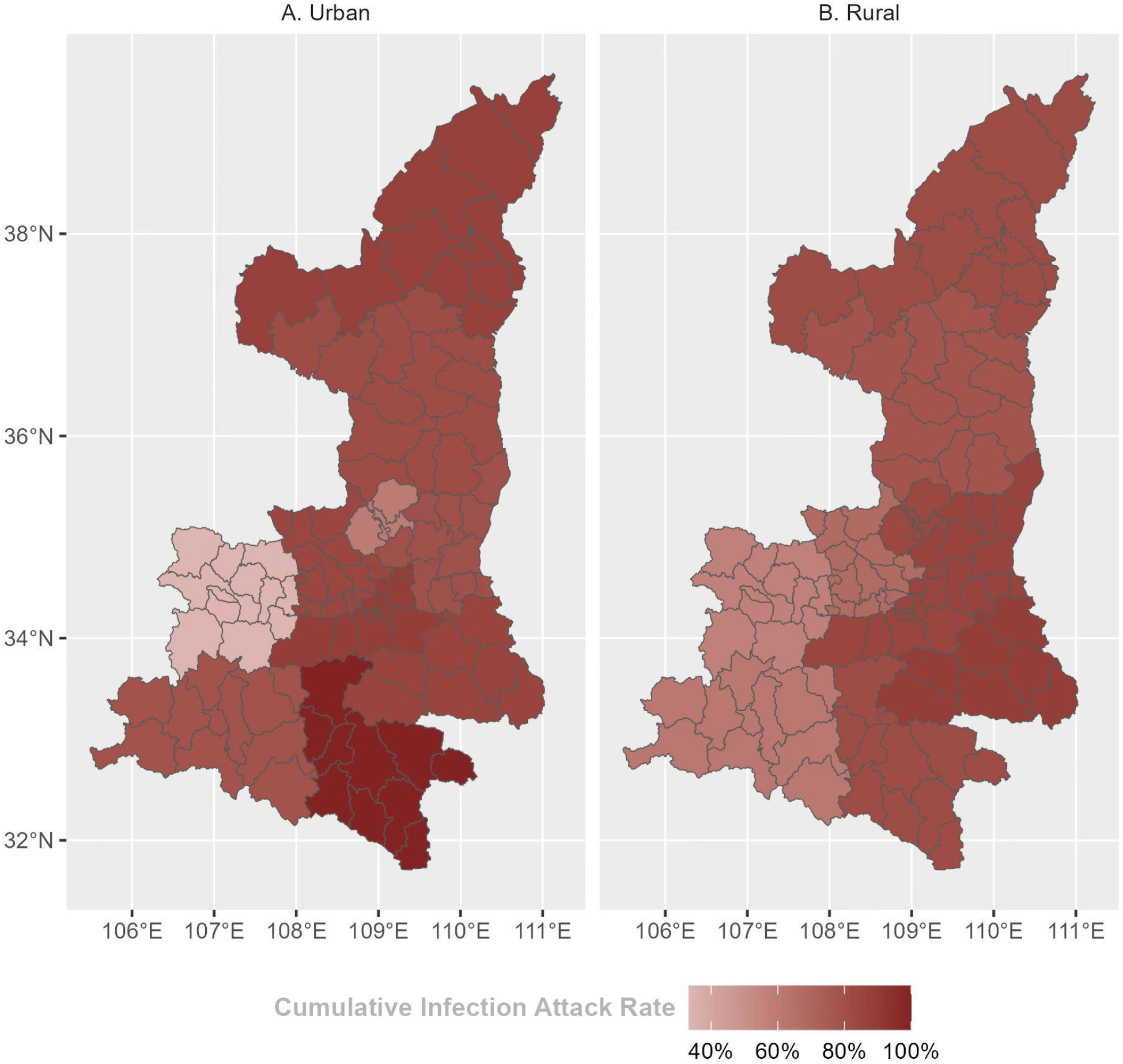
Time distribution of self-reported date of diagnosis for the COVID19 patients from Dec.1^st^, 2022 – Jan.15^th^ 2023. Left dotted line: The date when the 10 new COVID easing steps were announced (Dec. 7^th^, 2022). Right dotted line: The official beginning of the Spring Festival travel rush (Jan. 7^th^, 2023). A. The overall distribution of self-reported date of diagnosis for the COVID19 patients from Dec.1^st^, 2022 – Jan.15^th^ 2023. B. The distribution of self-reported date of diagnosis for the COVID19 patients from Dec.1^st^, 2022 – Jan.15^th^ 2023 grouped by types of residential address.

### Epidemiology characteristics of the returnees during the Spring Festival travel rush

A total of 271 returnees during the Spring Festival travel rush have provided specific dates of return. Among them, 229 individuals reported to be infected (84.5%). The majority of these patients (173, 75.5%) have the dates of self-reported diagnosis prior to their dates of return. The distributions of the self-reported date for diagnosis and the date of return for these patients were summarized in Figure 4. A steep rise in number of returnees was observed in Jan 13^th^ to 14^th^ 2023. The time distribution for the returnees’ self-reported diagnosis dates was similar to the overall transmission curves of COVID-19 outbreaks in Shaanxi Province.

**Figure 4.**
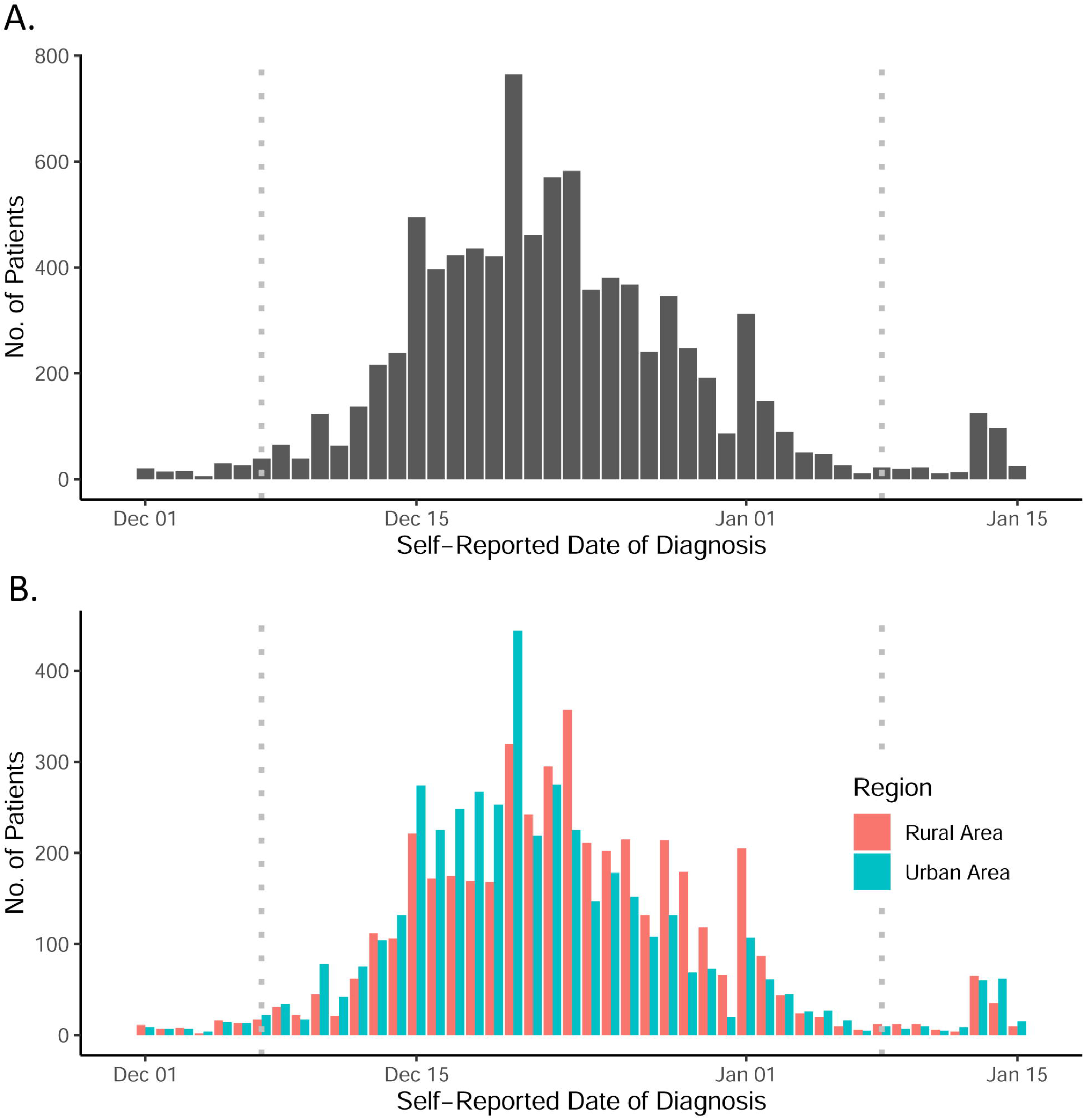
Time distribution for date of return and self-reported date of diagnosis for the COVID19 patients who returned their hometown for the Spring Festival from Dec.1^st^, 2022 – Jan.15^th^ 2023. Left dotted line: The date when the 10 new COVID easing steps were announced (Dec. 7^th^, 2022). Right dotted line: The official beginning of the Spring Festival travel rush (Jan. 7^th^, 2023).

### The pattern of COVID-19 symptoms reported by the study participants

The typical self-reported symptoms and proportions in COVID-19 patients were summarized in Table 2. The three most common symptoms were cough (5909, 57.6%), fever over 38□ (5707, 55.7%), and headache (4940, 48.2%). About 2.4% of the self-reported infected individuals (positive for rapid agent test or RT-PCR) have none of those typical symptoms. These test-positive individuals might be asymptomatic patients.

**Table 2.** Proportions of typical COVID19 symptoms reported by the patients.

| Symptoms | No.of Patients | Proportion in the Patients, % |
| --- | --- | --- |
| Cough | 5,909 | 57.6 |
| Fever over 38°C | 5,707 | 55.7 |
| Headache | 4,940 | 48.2 |
| Sore throat | 4,678 | 45.6 |
| Muscle or body aches | 4,286 | 41.8 |
| Congestion or runny nose | 3,224 | 31.4 |
| Throat itching | 2,648 | 25.8 |
| New loss of taste or smell | 1,465 | 14.3 |
| Chill | 1,088 | 10.6 |
| Nausea or vomiting | 778 | 7.6 |
| Red or sore eyes | 473 | 4.6 |
| Shortness of breath or difficulty breathing | 478 | 4.7 |
| None of the symptoms above | 244 | 2.4 |

## Discussion

The cumulative IAR of Shaanxi province is estimated to be 84.7% and this indicator in the urban areas is slightly higher compared to the rural areas in Shaanxi province. The peak of self-reported diagnosis for COVID-19 ranged from Dec. 15^th^, 2022 to Jan. 1^st^, 2023 at the provincial level. Compared to the rural areas, the COVID-19 outbreaks in the urban areas had a steep rise and decline. On the other hand, the peak of the epidemic in rural areas is a plateau which lasts for about 15 days. A small but steep rise between Jan 13^th^ to 14^th^, 2023 was identified indicating individuals spared from the first wave of the COVID-19 epidemic in Shaanxi province in Dec. 2022 might be endangered by the returnees during this Spring Festival.

The present study depicted the transmission dynamics patterns of the recent COVID-19 outbreaks in Shaanxi province based on the baseline data from a large scale cohort. Up to date, very few studies have been published to examine the transmission dynamics patterns of on-going COVID-19 epidemic in mainland China. A recent study conducted by Leung *et al*. estimated the transmission dynamics of SARS-CoV-2 Omicron BF.7 in Beijing after the adjustment of zero-COVID policy based on dynamic models [3].The cumulative IAR of Beijing is estimated to be 75.7% on Dec. 22^nd^ in that study, which is very similar to our current study in Shaanxi province despite the fact that the date of the diagnosis peak in Beijing is about 1 week earlier compared to our results of the urban areas in Shaanxi province. Beijing as a global fluent city and has become a central gateway connecting China and other parts of the world. It has higher risk level for the transmission of SARS-CoV-2 virus compared to Shaanxi province which is a landlocked province in northwest China.

An interesting observation is that although in general, based on data of the present study, the cumulative IAR was higher in the urban area compared to rural area in Shaanxi province (85.6% versus 83.7%), the rural area after removing those remote regions (defined by rural areas ≥10km from the nearby urban areas) has higher cumulative IAR compared to the urban area (92.9% versus 85.6%). One possible explanation for this finding is that rural areas which located near to the urban regions might have similar population density to the urban areas. It in turn makes these residents have comparable risk of exposure. In contrast to urban residents, residents of rural areas formed an “acquaintances society” in which people have more interpersonal communications with one another and are thus much more susceptible to infection than urban residents.

Besides the major peak of this round outbreaks (around Dec.20^th^, 2022), a small but steep rise for number of patients between Jan 13^th^ to 14^th^, 2023 was identified. The geographical distributions of the patients included in this small rise revealed that the patients were mostly identified in Shaanxi province’s core urban areas. It indicates that the effects of Spring Festival travel rush on transmission of COVID-19 might have emerged. Further examination of the patients of returnees revealed that, while the majority of these returnees (75.7%) were affected prior to their Spring Festival travels, many of them experienced symptom onset after returning hometown. The magnitude of this small increase is approximately 1/5 to 1/6 compared to the main peak of the epidemic in December 2022, implying that individuals spared from the first wave of COVID-19 outbreaks may be endangered by returnees during this Spring Festival.

In general, the symptom prevalence reported by patients in the current study is lower than in a previous study conducted by researchers in the United Kingdom. The top three symptoms in Omicron-infected people in that study were runny nose (76.5%), headache (74.7%), and sore throat (70.5%). In the current study, however, the prevalence of these symptoms decreased by about 20-30% in the patients [9-11]. The various Omicron variants could explain at least some of the difference in symptom prevalence. Furthermore, different genetic backgrounds and characteristics of the study populations could be a contributing factor to this disparity. Furthermore, the proportion of asymptomatic patients reported in the current study is quite low when compared to other reports [12-13]. Because active screening using RT-PCR tests was not performed for study participants in this study, the proportion of asymptomatic patients may be underestimated.

This study suffered from several limitations. Firstly, as an observational study, it suffers from several biases such as sampling bias and recall bias, which may affect the results. In addition, the numbers of samples from the ten cities were not balanced, which could have an impact on the accuracy of the transmission dynamics curves. The self-reported date of diagnosis might be inaccurate in depicting the transmission patterns of the epidemic.

## Conclusion

According to the COVID-19 cumulative IAR data, the herd community was primarily achieved in Shaanxi province’s urban and rural areas. The epidemics in other provinces might be similar to that of Shaanxi despite that there are differences of detail in transmission dynamics. The epidemic in Shaanxi province has been exacerbated by mass population movement during the Spring Festival travel rush in both urban and rural areas. In future, surveillance should be performed to monitor the spread of SARS-CoV-2 infections and track relevant epidemiological indicators in Shaanxi province.

## Data Availability

The data that support the findings of this study are available on request from the corresponding author QW. The data are not publicly available due to local laws, regulations and restrictions.

## Acknowledgement

We want to thank the student research volunteers for their focusing on work during winter vacations. We also want to thank the study participants. Without them no meaningful research could be done.

## Authors Contributions

BM proposed the first idea of performing this survey. QW, MS, SZ, and YZ then thoroughly discussed the idea and make it a completed study design. BM directed the large scale field work. YN, SC, JL, HH, ZW, and LL contributed significantly in the field work and data collection. TZ, BM and MS performed data analyses. TZ, QW, and BM drafted the first version of manuscript. All the authors contributed to the revisions of the manuscript. All authors have seen and approved the manuscript.

## Reference

1. Xinhua. China Focus: China releases measures to optimize COVID-19 response, <https://english.news.cn/20221111/d4399114a082438eaac32d08a02bf58d/c.html> (2022).

2. Xinhua. China Focus: COVID-19 response further optimized with 10 new measures, <https://english.news.cn/20221207/ca014c043bf24728b8dcbc0198565fdf/c.html> (2022).

3. Leung K, Lau EHY, Wong CKH, et al. Estimating the transmission dynamics of SARS-CoV-2 Omicron BF.7 in Beijing after the adjustment of zero-COVID policy in November - December 2022. Nature Medicine. 2023. [print ahead of publications]

4. Wang X, Ji X. Sample Size Estimation in Clinical Research: From Randomized Controlled Trials to Observational Studies. Chest. 2020;158(1S):S12–S20.

5. PA Harris, R Taylor, R Thielke, et al. Research electronic data capture (REDCap) – A metadata-driven methodology and workflow process for providing translational research informatics support, J Biomed Inform. 2009;42(2):377–81.

6. PA Harris, R Taylor, BL Minor, et al. The REDCap consortium: Building an international community of software partners, J Biomed Inform. 2019;95:103208.

7. Gao XY, Mi BB, Wu WT,et al [Application of electronic data acquisition system REDCap in large natural population-based cohort studies]. Zhonghua Liu Xing Bing Xue Za Zhi. 2020;41(9):1542-1549. In Chinese.

8. R Core Team. R: A language and environment for statistical computing. R Foundation for Statistical Computing, Vienna, Austria. 2022. URL https://www.R-project.org/.

9. Espenhain L, Funk T, Overvad M, et al. Epidemiological characterisation of the first 785 SARS-CoV-2 Omicron variant cases in Denmark, December 2021. Euro Surveill. 2021;26(50):2101146.

10. Menni C, Valdes AM, Polidori L, et al. Symptom prevalence, duration, and risk of hospital admission in individuals infected with SARS-CoV-2 during periods of omicron and delta variant dominance: a prospective observational study from the ZOE COVID Study. Lancet. 2022;399(10335):1618–1624.

11. Marquez C, Kerkhoff AD, Schrom J, et al. COVID-19 Symptoms and Duration of Rapid Antigen Test Positivity at a Community Testing and Surveillance Site During Pre-Delta, Delta, and Omicron BA.1 Periods. JAMA Netw Open. 2022;5(10):e2235844.

12. Garrett N, Tapley A, Andriesen J, et al. High Rate of Asymptomatic Carriage Associated with Variant Strain Omicron. medRxiv [Preprint]. 2022; 14:2021.12.20.21268130.

13. Yu W, Guo Y, Zhang S, et al. Proportion of asymptomatic infection and nonsevere disease caused by SARS-CoV-2 Omicron variant: A systematic review and analysis. J Med Virol. 2022;94(12):5790–5801.

