## Supplementary Materials for "Exploring the Transmission Dynamics of the COVID-19 Outbreaks after Dec. 2022 in Shaanxi Province, China: Analysis of Baseline Data from A Large Scale Cohort"

**Supplementary Tables.**

**Supplementary Table S1.** The cumulative infection rate in different cities of Shaanxi province.

| City | Urban areas | | | Rural areas | | |
| --- | --- | --- | --- | --- | --- | --- |
|  | No. of infected  people | No. of study  participants | Cumulative infection  attack rate（%） | No. of infected  people | No. of study  participants | Cumulative infection  attack rate（%） |
| Xi'an | 2,537 | 2,865 | 88.6 | 1,216 | 1,432 | 84.9 |
| Yulin | 488 | 560 | 87.1 | 285 | 348 | 81.9 |
| Tongchuan | 3 | 5 | 60.0 | 157 | 188 | 83.5 |
| Ankang | 7 | 7 | 100.0 | 580 | 709 | 81.8 |
| Weinan | 653 | 814 | 80.2 | 2,592 | 3,005 | 86.3 |
| Baoji | 1 | 3 | 33.3 | 114 | 199 | 57.3 |
| Yan'an | 390 | 475 | 82.1 | 135 | 173 | 78.0 |
| Xianyang | 665 | 786 | 84.6 | 23 | 34 | 67.7 |
| Shangluo | 53 | 62 | 85.5 | 70 | 79 | 88.6 |
| Hanzhong | 263 | 333 | 79.0 | 21 | 34 | 61.8 |
| Total | 5,060 | 5,910 | 85.6 | 5,193 | 6,201 | 83.7 |

**Supplementary Table S2.** Number of reported patients in different cities and types of residential address between Jan. 13^th^ and Jan 14^th^, 2023.

| City | Urban areas | Rural areas |
| --- | --- | --- |
| Xi’an | 51 | 20 |
| Weinan | 31 | 27 |
| Yan’an | 5 | 7 |
| Yulin | 16 | 14 |
| Xianyang | 26 | 2 |
| Shangluo | 2 | 3 |
| Ankang | 0 | 6 |
| Tongchuan | 0 | 5 |
| Baoji | 0 | 2 |
| Hanzhong | 8 | 4 |
| Total | 139 | 90 |

**Supplementary Figures**

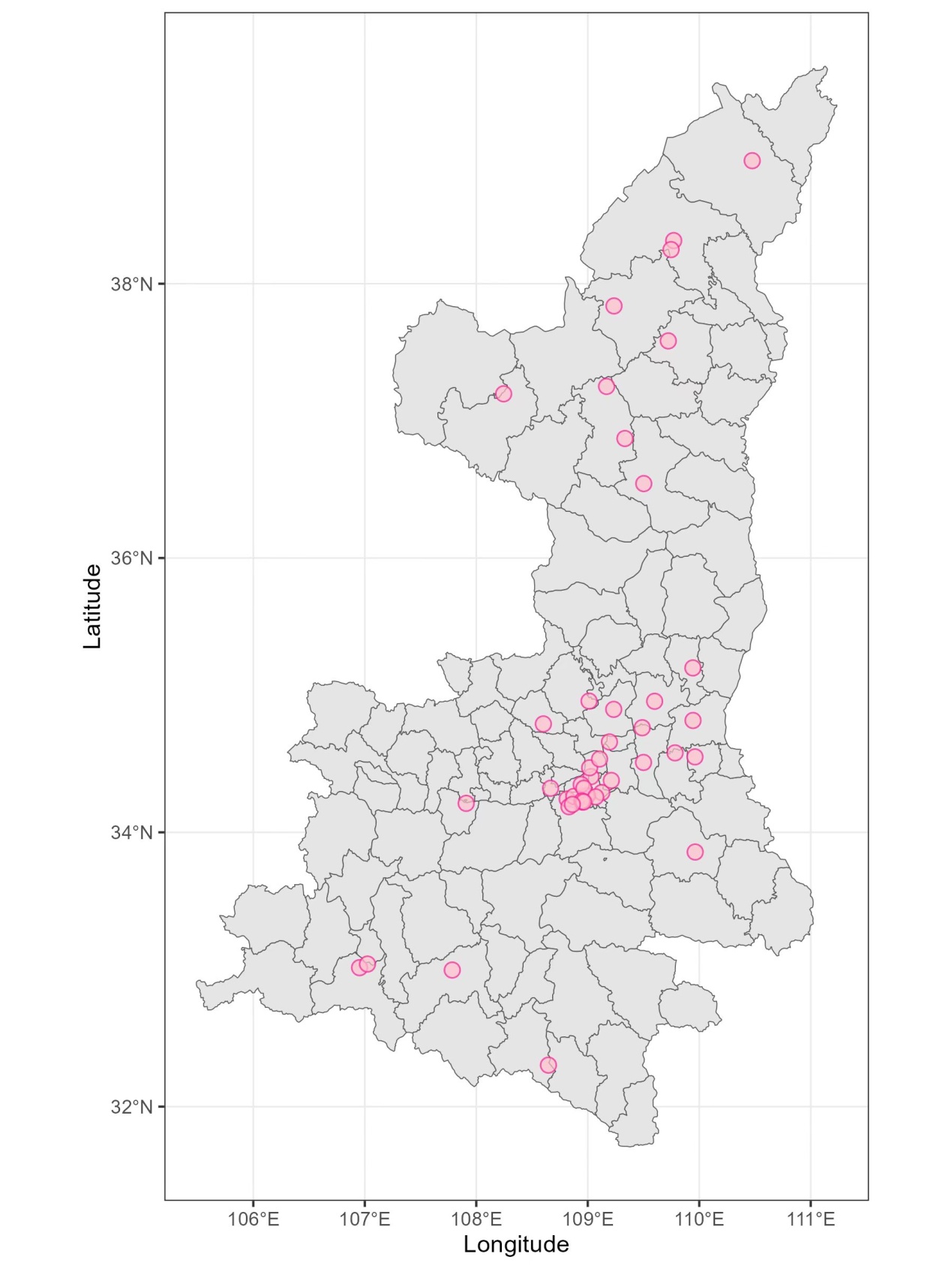

**Supplementary Figure S1.** Sampling sites in Shaanxi province

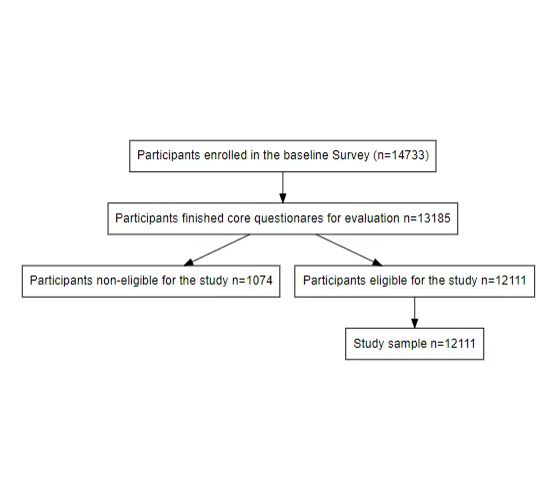

**Supplementary Figure S2.** Flow chart of the study participants enrollment.

**A sample of the survey questionnaire**

**COVID-19 Surveillance In Urban And Rural Areas Of Shaanxi Province(Baseline Survey)**

Record ID：

**1.** Mobile phone number:

**2.** Name:

**3.** ID number:

**4.** Current age (years):

**5.** Gender:

**6.** Current residence:

**7.** What is your type of residence?

 Urban area

 Villages and towns

**8.** What is your usual residence?

 This village/community

 Outside village/outside community in this county

 Counties outside the city

 City outside the province of shaanxi

 Provincial

 Overseas

**9.** How many people live in your family (besides you)?

**10.** How many people in your family are currently infected with COVID-19 (besides you)?

**11.** Are you the head of the household?

 Yes

 No

**11.1** Name of householder:

**12.** Do you have any of the following underlying medical conditions (select at least 1)?

 Hypertension

 Diabetes

 Chronic Obstructive Pulmonary Disease

 Chronic Bronchitis

 Asthma

 CVD(Cardiovascular Disease)

 Stroke

 Chronic Nephrosis

 Chronic Liver Disease

 Hematological System Diseases

 Disease Of Immune System

 Malignant Tumor

 Other Underlying Diseases

 None Of The Above

**13.** Are you infected with COVID-19?

 Never infected

 Infected with once

 Infected with two or more

**14.** What was your basis for confirming the infection?

 ConsciouslyAsymptomatic

 Self-evident symptom

 Antigen test positive

 Nucleic acid test positive

**14.1** The date of your antigen positive test is:

**14.2** The date of your positive nucleic acid test is:

**15.** During infection, do you have the following symptoms?

 Body temperature over 38.0℃:

 Cough (if you have a basis for coughing, more severe than usual)

 Muscle pain (if you usually have muscle pain, is it worse?)

 Snotty

 [Headache](javascript:;)

 Sore throat

 Itchy throat

 Nausea, vomiting, or diarrhea

 Polypnea

 Unable to taste or smell

 Red or sore eyes

 Shiver

 None of the above

16. What is the degree of pain all over?

 Mild pain

The pain is moderate and tolerable

 Unbearable pain

17. Have you sought medical attention after infection?

 Outpatient/emergency

 In hospital

 Not seen a doctor

18. Do you still have symptoms?

 Yes, it's still serious

 Yes, but it's getting better

 No

 19. Have you been tested for antigens in the last 48 hours?

 Yes, negative

 Yes, positive

 No

20. Have you taken a nucleic acid test in the last 48 hours?

 Yes, negative

 Yes, positive

 No

21. In the past 48 hours, have you participated in any gathering activities (five or more people in a confined indoor environment)?

 Yes

 No

22. How many people have you had close contact with in the last 48 hours?

23. What medical drugs do you have at present?

 Antigen detection reagent

 Febrifuge

 Pectoral

 Oximetry equipment (e.g. Clip-on oximeter)

 Breathing machine

 Oxygenerator

 Other

 None of the above

24. What is the source of all your medical drugs?

 Household supplies

 Purchase by oneself

 Official or unofficial channels for mutual assistance

 Other

25. What is your preferred means of transportation at present?

 Walk

 Bicycle

 Electromobile

 Motorbike

 Subway

 Bus

 Private car

 Other

26. Do you have any plans to visit relatives across provinces/cities during the Spring Festival?

 Yes, interprovincial

 Yes, within the province and across the city

 Yes, the city is cross-county

 Yes, county flow

 No

**First follow-up**

Record ID：

1. Have any relatives returned since the last survey?

 Yes

 No

2. Have you been infected with COVID-19 since the last survey?

 Uninfected

 Infected

3. Do you still have symptoms?

 Yes, it's still serious

 Yes, but it's getting better

 No

4. Have you been tested for antigens since the last survey?

 Yes, negative

 Yes, positive

 No

5. Have you taken a nucleic acid test since the last investigation?

 Yes, negative

 Yes, positive

 No

6. Since the last survey, have you participated in any gathering activities (more than 5 people in a confined indoor environment)?

 Yes

 No

7. How many people have you had close contact with since your last survey?

8. What medical drugs do you have at present?

 Antigen detection reagent

 Febrifuge

 Pectoral

 Oximetry equipment (e.g. Clip-on oximeter)

 Breathing machine

 Oxygenerator

 Other

 None of the above

9. What is the source of all your medical drugs?

 Household supplies

 Purchase by oneself

 Official or unofficial channels for mutual assistance

 Other

**Second follow-up**

Record ID：

1. Have any relatives returned since the last survey?

 Yes

 No

2. Have you been infected with COVID-19 since the last survey?

 Uninfected

 Infected

3. Do you still have symptoms?

 Yes, it's still serious

 Yes, but it's getting better

 No

4. Have you been tested for antigens since the last survey?

 Yes, negative

 Yes, positive

 No

5. Have you taken a nucleic acid test since the last investigation?

 Yes, negative

 Yes, positive

 No

6. Since the last survey, have you participated in any gathering activities (more than 5 people in a confined indoor environment)?

 Yes

 No

7. How many people have you had close contact with since your last survey?

8. What medical drugs do you have at present?

 Antigen detection reagent

 Febrifuge

 Pectoral

 Oximetry equipment (e.g. Clip-on oximeter)

 Breathing machine

 Oxygenerator

 Other

 None of the above

9. What is the source of all your medical drugs?

 Household supplies

 Purchase by oneself

 Official or unofficial channels for mutual assistance

 Other
